# Reduced odor detection and hedonic changes in asymptomatic university students as SARS-CoV-2 emerged locally

**DOI:** 10.1101/2020.06.17.20106302

**Authors:** Julie Walsh-Messinger, Sahar Kaouk, Hannah Manis, Rachel Kaye, Guillermo Cecchi, Pablo Meyer, Dolores Malaspina

**Affiliations:** Department of Psychology, University of Dayton, Dayton, OH; Department of Psychiatry, Wright State University Boonshoft School of Medicine, Dayton, OH; Department of Otolaryngology-- Head & Neck Surgery, Rutgers New Jersey Medical Center, Newark, NJ; IBM Thomas J. Watson Research Center, Yorktown Heights, NY; Departments of Psychiatry, Neuroscience, and Genetics and Genomics, Icahn School of Medicine at Mount Sinai, New York, NY

**Keywords:** Olfaction, SARS-CoV-2, COVID-19, anosmia, hyposmia

## Abstract

Aerosol droplets have emerged as the primary mode of SARS-Cov-2 transmission and can be spread by infectious asymptomatic/pre-symptomatic persons rendering indicators of latent viral infection essential. Olfactory impairment is now a recognized symptom of COVID-19 and is rapidly becoming one of the most reliable indicators of the disease. We compared olfaction data from asymptomatic students, who were assessed as SARS-CoV-2 was unknowingly spreading locally, to students tested prior to the arrival of the virus. This study was naturalistic by design as testing occurred in the context of four research studies, all of which used the same inclusion/exclusion criteria and the same protocol to objectively assess odor detection, identification, and hedonics with physiological tests. Data from students (Cohort II; N=22) with probable SARS-CoV-2 exposure were compared to students tested just prior to local virus transmission (Cohort I; N=25), and a normative sample of students assessed over the previous four years (N=272). Students in Cohort II demonstrated significantly reduced odor detection sensitivity compared to students in Cohort I (t=2.60; P=.01; d=0.77; CI, 0.17, 1.36), with a distribution skewed towards reduced detection sensitivity (D=0.38; P=.005). Categorically, the exposed group was significantly more likely to have hyposmia (OR=7.74; CI, 3.1, 19.40), particularly the subgroup assessed in the final week before campus closure (OR=13.61; CI, 3.40, 35.66;). The exposed cohort also rated odors as less unpleasant (P<.001, CLES=0.77). A limitation of our study is that participants were not tested for COVID-19 as testing was unavailable in the area. Objective measures of olfaction may detect olfactory impairment in asymptomatic persons who are otherwise unaware of smell loss. The development of cost-effective, objective olfaction tests that could be self-administered regularly could aid in early detection of SARS-CoV-2 exposure, which is vital to combatting this pandemic.

## INTRODUCTION

Coronaviruses (CoVs) are a group of enveloped single stranded RNA viruses associated with the common cold in adult humans and upper respiratory viruses in other mammals and birds [1]. The viruses were considered to produce mild symptoms until the highly lethal SARS-Cov-1 presented in 2003, followed by Middle East respiratory syndrome (MERS-CoV) in 2012. The novel SARS-CoV-2 virus emerged in Wuhan in 2019 and has rapidly became a worldwide pandemic, challenging health services and economic stability worldwide.

Self-reports of diminished smell (hyposmia/anosmia) and taste (ageusia/dysguesia) led to the inclusion of chemosensory impairment as recognized COVID-19 symptoms by the World Health Organization (WHO) and Center for Disease Control (CDC) [2], fueling efforts to identify the pathobiology, particularly for the olfactory symptoms [3]. While the proportion of cases with olfactory complaints varies by assessment approach and with sample age and sex [4], there is a consensus that olfactory symptoms can present early in the course of the infection, sometimes as the only symptom [5], and appears to be a better predictor of COVID-19 positivity than other symptoms such as fever and cough [6]. Early olfactory impairment may be associated with better outcomes [7], although a recent report suggests that there may be a subgroup with COVID-19 smell loss that experiences longer duration of symptoms [6].

Early research findings suggest that SARS-Cov-2 may primarily spread through aerosol droplets [8]. which can be respired by asymptomatic/pre-symptomatic persons [9]; thus, indicators of latent infection are of crucial interest. Entry of SARS-CoV-2 into cells involves spike (S) protein binding to angiotensin-converting enzyme 2 (ACE2) along with host proteases, principally transmembrane serine protease 2 (TMPRSS2). Olfactory receptor neurons do not express these receptors [10], so the earliest olfactory changes likely reflect non-neuronal olfactory epithelial cell entry, including cells that initiate ultra-rapid immune responses [11]. The nasal epithelium contains innate immune-associated tissue, the nasal-associated lymphoid tissue. This phylogenetically ancient system is the first line of immune defense against inspired or ingested viruses and other pathogens, organizing systemic inflammatory processes [12] that may be protective against more severe pulmonary involvement. As diminished odor sensitivity in rhinosinusitis is correlated with cytokine responses [13, 14], diminished sense of smell may not entail overt pathology to olfactory receptor neurons.

Discerning the mechanism for the earliest loss of olfactory sensitivity may be a key to early detection of SARS-CoV-2 exposure and infection, although prospective studies on the earliest olfactory impairment is limited. Most studies have relied on self-reported symptoms, which are significantly less sensitive to dysfunction than standard olfactory measures. To date, no studies have comprehensively assessed odor detection, identification, and hedonic judgments in asymptomatic and pre-symptomatic persons exposed to COVID-19. Odor detection and identification involve different olfactory pathways, but to our knowledge, they have not yet been distinguished in the COVID-19 literature. Additionally, no studies have compared olfactory function in exposed persons to a large normative sample of age-appropriate and comorbidity-matched participants, nor had the availability of comparison measures from a cohort tested prior to the emergence of the pandemic.

We have applied our unique data, drawn from four years of systematic olfactory function assessments in healthy undergraduate students, to address these significant gaps in the literature. Data from an asymptomatic cohort of students exposed to SARS-CoV-2 as it became endemic were compared to data from a cohort tested in the weeks prior to the arrival of the virus and to ecologically valid comparison groups.

## METHOD

### Participants

#### Olfaction Tested Cohorts

Undergraduate students (N=367) participated in one of four research studies of odor detection sensitivity and identification conducted at a private Midwestern university from 2016–2020. The studies all followed the same olfaction testing protocol and all participants were asked to not wear perfume, cologne or strongly scented lotions, or smoke or vape at least four hours prior to their appointment or consume food or beverages 15 minutes prior to testing. The studies were all approved by the university’s Research Review and Ethics Committee, a subcommittee of the Institutional Review Board. All participants provided voluntary written informed consent, obtained in accordance with the Declaration of Helsinki, and earned research experience class credit for their participation.

We retrospectively separated participants into cohorts (Fig 1a) based on campus exposures to SARS-CoV-2, exclusive of 64 students who reported or exhibited nasal congestion, allergic rhinitis, cough, or other respiratory symptoms (N=50) or smoked cigarettes or vaped (N=14) within four hours of testing. Students (Cohort IV) who were tested February 14–26, 2020 (N=9) when COVID-19 exposure is uncertain were only included in analyses where noted. Cohort I (N=25) was comprised of those tested before local transmission of the virus, from January to early February 2020. Cohort II (N=22) included students tested from late February to early March 2020 which corresponds to an observed viral outbreak on campus. A subset of this group (Cohort IIa) was tested immediately before social distancing was enforced on campus. Publicly available health records confirm that SARS-CoV-2 was endemic to the area as early as January 13, 2020 and became more widespread by the end of February [15].

**Figure 1.**
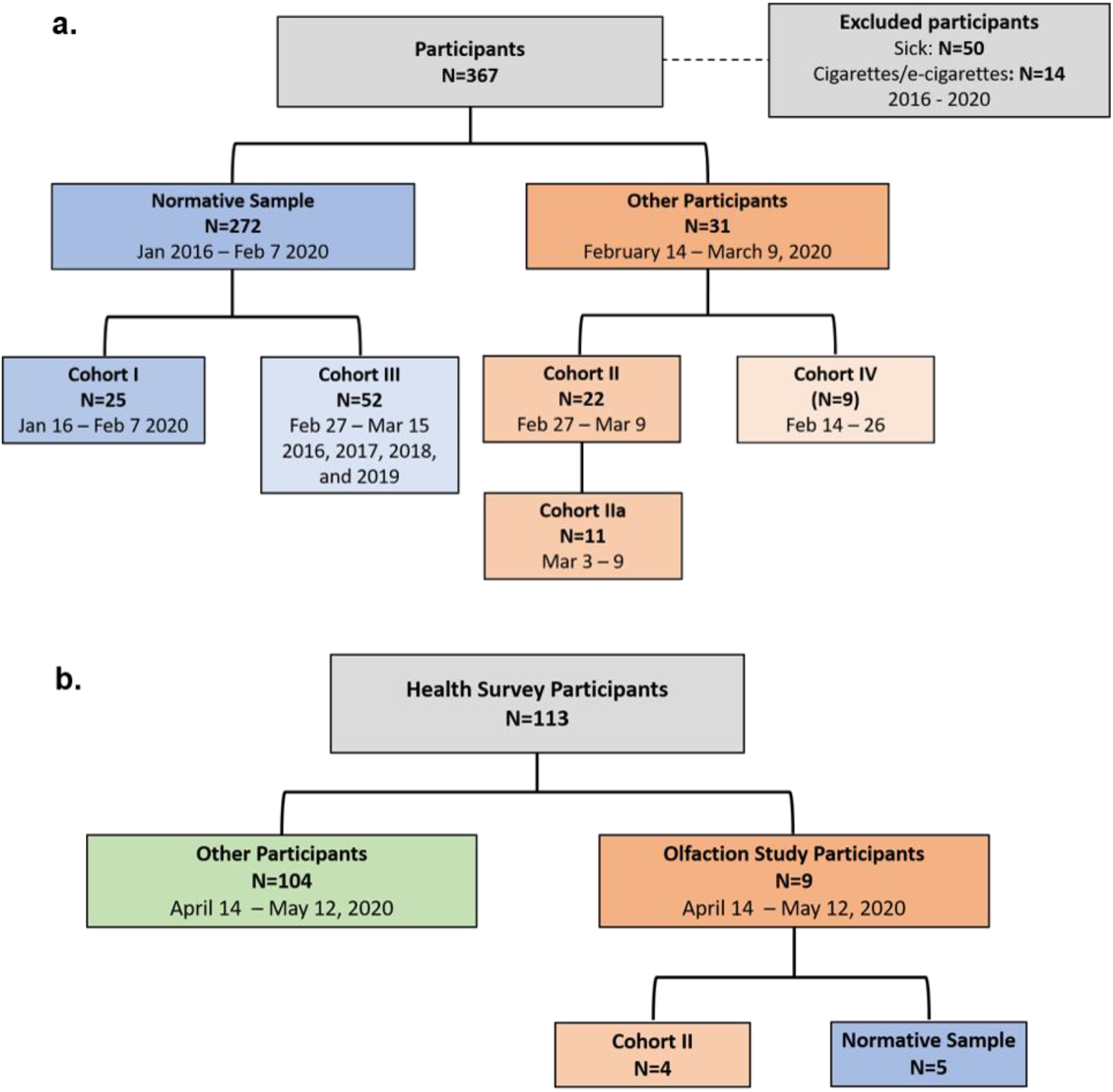
Sample sizes and assessment time periods for **a)** olfaction tested cohorts, and **b)** health survey participants.

To assess the potential impact of seasonal factors, comparisons were made to the students tested late February and early March 2016-2019 (Cohort III; N=52). Normative performance was determined from an ecologically valid comparison group of all students (N=272) tested in this laboratory prior to February 7, 2020. The cohorts did not differ in age, sex, or cigarette use (see Table 1).

**Table 1.** Demographic information for all tested cohorts.

|  | Cohort II | Normative Sample |  |  | Cohort II vs Cohort I |  | Cohort II vs Cohort III |  | Cohort II vs Normative Sample |  |
| --- | --- | --- | --- | --- | --- | --- | --- | --- | --- | --- |
|  | N=22 | All‡<br>N=272 | Cohort I<br>N=25 | Cohort III†<br>N=52 | t | p | U | p | D | p |
| Age, Mean (SD) | 19.45 (1.23) | 19.28 (1.11) | 19.08 (1.08) | 19.31 (1.09) | 0.93 | .36 | 550.50 | .79 | 1.92 | .83 |
| Menstrual cycle day, Mean (SD)§ | 15.41 (8.56) | 17.31(8.20) | 14.59 (8.51) |  | -0.97 | .35 |  |  |  |  |
|  |  |  |  |  | **X²** | **p** | **X²** | **p** | **X²** | **p** |
| Sex — number (%) |  |  |  |  | 0.00 | .99 | 0.482 | .49 | 0.49 | .83 |
| Male | 7 (32) | 83 (34) | 8 (32) | 21 (40) |  |  |  |  |  |  |
| Female | 15 (68) | 160 (66) | 17 (68) | 31 (60) |  |  |  |  |  |  |
| Racial/Ethnicity — number (%) |  |  |  |  | 0.41 | 0.52 |  |  |  |  |
| Asian or Asian American | 1 (4.5) | (12) | 1 (4) |  |  |  |  |  |  |  |
| Black or African American | 2 (9) | (3) | 3 (12) |  |  |  |  |  |  |  |
| Hispanic White | 2 (9) | (12) | 4 (16) |  |  |  |  |  |  |  |
| Non-Hispanic White | 16 (73) | (71) | 16 (64) |  |  |  |  |  |  |  |
| Other | 1 (4.5) | (3) | 1 (4) |  |  |  |  |  |  |  |
| Cigarette Use N (%) |  |  |  |  | 0.39 | .53 | 0.64 | .43 | 0.94 | .33 |
| None | 19 (86) | 250 (92) | 22 (88) | 48 (92) |  |  |  |  |  |  |
| Yes, but not daily | 1 (5) | 14 (5) | 1 (4) | 3 (6) |  |  |  |  |  |  |
| Yes, daily | 2 (5) | 4 (2) | 2 (8) | 1 (2) |  |  |  |  |  |  |
| Use of Vaping Devices N (%) |  |  |  |  | 2.02 | 0.37 |  |  |  |  |
| None | 17 (77) | (82) | 18 (72) |  |  |  |  |  |  |  |
| Yes, but not daily | 3 (14) | (14) | 2 (8) |  |  |  |  |  |  |  |
| Yes, daily | 2 (9) | (3) | 5 (20) |  |  |  |  |  |  |  |

Tests statistics refer to: t=Independent Samples t-test; U=Mann-Whitney U-test; D=Two Samples Kolmogorov–Smirnov test
§Females only; converted to a 28-day cycle; Cohort I and II N=15 females; data missing from Cohort I females (N=2)
‡Data missing: race/ethnicity and use of vaping devices (N=209); menstrual cycle in females (N=137); Counts were not reported and comparative
analyses were not conducted due to the large percentage of missing data.
†Data missing: race/ethnicity and use of vaping devices (N=47); menstrual cycle in females (N=26); Counts and percentages were not reported and comparative analyses were not conducted due to the large percentage of missing data.

#### Health Survey Sample

Separate Research Review and Ethics Committee approval was obtained to conduct an online health survey following reports that anosmia/hyposmia was potentially linked to COVID-19. Of the 130 students who signed-up for the survey to earn research experience credit for class, 113 (87%) provided written informed consent and subsequently completed the survey administered via Qualtrics (Qualtrics^XM^, Provo, UT) (see Fig 1b). A subset of students from the aforementioned cohorts completed this survey (N=9), including four students from Cohort II, and provided consent to link their survey and olfaction data.

### Measures

#### Olfaction

All measures were administered by systematically trained undergraduate and graduate research assistants. The *Sniffin’ Sticks n-Butanol Threshold Test* [16] assessed odor detection sensitivity using a single-staircase forced-choice design. The test utilizes pen-like odor dispensing devices, 32 of which are blanks (no odor) and 16 of which contain varying concentrations of n-butanol (alcohol). The test employs a single-staircase forced-choice design in which the test administrator presents odor triplets directly below the blindfolded participant’s nostril (two blanks, one with n-butanol) one at a time in random order for five seconds each. After each triplet presentation, the participant was asked to identify the pen that contained the odor. Scores range from 0 (low) to 16 (high). Participants were classified as anosmic, hyposmic, or normosmic based on published data [16].

The *Sniffin’ Sticks Identification Test* [16], is comprised of 16 pen-like devices that deliver common odorants (e.g. orange, garlic, fish, etc.). The test administrator presented each odor pen directly under the participant’s nostril, one at a time for 3-4 seconds. After each presentation, the test administrator read four response choices and the participant choose the one that best described the odor. Scores range from 0 (low) to 16 (high). Participants then rated each the pleasantness and unpleasantness of each odor on two five-point Likert scales ranging from “0” (not pleasant/unpleasant) to “4” (extremely pleasant/unpleasant). Responses across all 16 odors were summed to yield total pleasantness and unpleasantness scores, with scores for each scale ranging from “0” (not pleasant/unpleasant) to 64 (extremely pleasant/unpleasant).

#### Health Survey

Participants were surveyed between April 14^th^ and May 10^th^, 2020 as to whether they had experienced fever, sore throat, lymphadenopathy, cough, dyspnea, nasal congestion, myalgias, fatigue, diarrhea, and/or emesis (excluding alcohol related incidents) since January 1, 2020. For each symptom endorsed, follow-up queries assessed severity, timing, and frequency. Additional questions asked chronic health conditions and whether they suspected or formally diagnosed with infectious diseases (influenza, COVID-19, strep pharyngitis, mononucleosis, and/or common cold) during this period, followed-up with questions regarding test results (if any), timing, concurrent symptoms, and

A modified version of the *Self-Report Inventory to Measure Olfactory Dysfunction and Quality of Life* [17], assessed changes in the sense of smell during a one-month period and the impact of any change on daily functions, modified to also assess changes in taste, including sweet, salty, bitter, and sour. Participants were asked whether they experienced any changes in their sense of smell or taste during January, February, March, or April 2020; if yes, they were asked to complete the measure for that month.

#### Data analysis

Data analyses were conducted with SPSS 23.0 (IBM, Armonk, NY). Participants with missing data were excluded from analyses, as detailed in the legends of Tables 1 and 2. With the exception of Cohort IIA’s odor pleasantness ratings, all data was normally distributed. Independent sample t-tests and chi-squares were used to test for cohort differences in demographic characteristics. Independent sample t-tests were employed to compare the olfaction performance of Cohorts I and II. Due to unequal sample sizes Mann-Whitney U tests were used to compare the olfaction performance of Cohort IIa to Cohorts I and III and to compare Cohort II to Cohort III.

To ensure that potential group differences were not the result of anomalous Cohort I data, a two-sample Kolmogorov-Smirnov (K-S) test was used to compare the frequency distributions of Cohorts II and IIa to the normative sample distribution. One-sample t-tests were used to compare the exposed cohorts to published norms. Cohen’s d, Common Language Effect Size (CLES) and 95% confidence intervals [CI] were computed using the Psycometrica online calculator [18]. Odds ratios were employed to compare risk of hyposmia in the weeks leading up to the closure of campus to the normative sample.

## RESULTS

### Odor Detection

Means and standard deviations for the olfaction measures are presented in Table 2. Significantly reduced odor detection sensitivity was demonstrated by Cohort II, the exposed cohort, compared to Cohort I, which was assessed prior to SARS-CoV-2’s emergence in the area (t=2.60; P=.01; d=0.77; CI, 0.17, 1.36). Cohort IIa, the subsample tested in the final week before social isolation, was also significantly impaired (U=58.50; P=.007; CLES=0.77; CI, 0.31, 1.81). Comparisons of odor sensitivity thresholds to those of students during the same time period in the previous four years (Cohort III) confirmed the absence of a seasonal explanation, with Cohort IIa showing less odor detection sensitivity (U=162.50; P=.03; CLES=0.75; CI, 0.30, 1.64), and a similar trend for the entire Cohort II (U=411.50; P=.06; CLES=0.63; CI=-.06, 0.95). A comparison of the distribution of Cohorts II and IIa to the normative sample distribution showed significant rightward (positive) skewing, indicating reduced odor detection sensitivity for Cohorts II (D=0.38; P=.005; Fig 2) and IIa (D=0.52, P=.007). Similarly, Cohorts II (t=-2.92., P=.008) and IIa (t=-3.39 P=.007) had diminished odor detection in comparison to published normative data [16].

**Table 2.** Medians, Means, standard deviations, and diagnostic classification for olfaction measures, by cohort.

|  | Cohort II |  | Normative Sample |  |  |
| --- | --- | --- | --- | --- | --- |
|  | All | Cohort IIa | All | Cohort I | Cohort III |
|  | N=22 | N=11 | N = 272 | N=25 | N=52 |
| Detection Sensitivity |  |  |  |  |  |
| Mean (SD) | 7.32 (3.21) | 6.50 (2.76) | 8.40 (2.15) | 9.79 (3.24) | 8.26 (19.31) |
| Median (IQR) | 6.38 (5.31) | 5.75 (5.25) | 8.38 (2.50) | 10.00 (4.88) | 8.50 (2.0) |
| % Correctly Identified |  |  |  |  |  |
| Mean (SD) | 72.44 (12.74) | 67.05 (11.21) | 71.22 (10.26) | 71.25 (11.98) | 72.36 (9.37) |
| Median (IQR) | 75.00 (19.00) | 68.75 (19.00) | 72.00 (13.00) | 75.00 (9.00) | 75.00 (12.60) |
| Pleasantness Ratings |  |  |  |  |  |
| Mean (SD) | 25.10 (7.86) | 23.20 (9.03) | 24.55 (7.23) | 25.20 (7.62) | 24.79 (7.77) |
| Median (IQR) | 24.00 (11.00) | 20.00 (8.00) | 24.00 (11.00) | 14.00 (13.00) | 24.00 (12.75) |
| Unpleasantness Ratings |  |  |  |  |  |
| Mean (SD) | 12.05 (6.18) | 13.55 (5.99) | 18.54 (7.42) | 14.52 (4.07) | 19.37 (7.39) |
| Median (IQR) | 10.00 (10.75) | 12.00 (10.00) | 18.00 (10.00) | 14.00 (6.00) | 19.00 (11.75) |
| Smell Diagnosis — no. (%) |  |  |  |  |  |
| Hyposmia | 11 (50) | 7 (64) | 31 (11) | 2 (8) | 2 (4) |
| Normosmia | 11 (50) | 4 (36) | 241 (89) | 23 (92) | 44 (96) |

**Figure 2:**
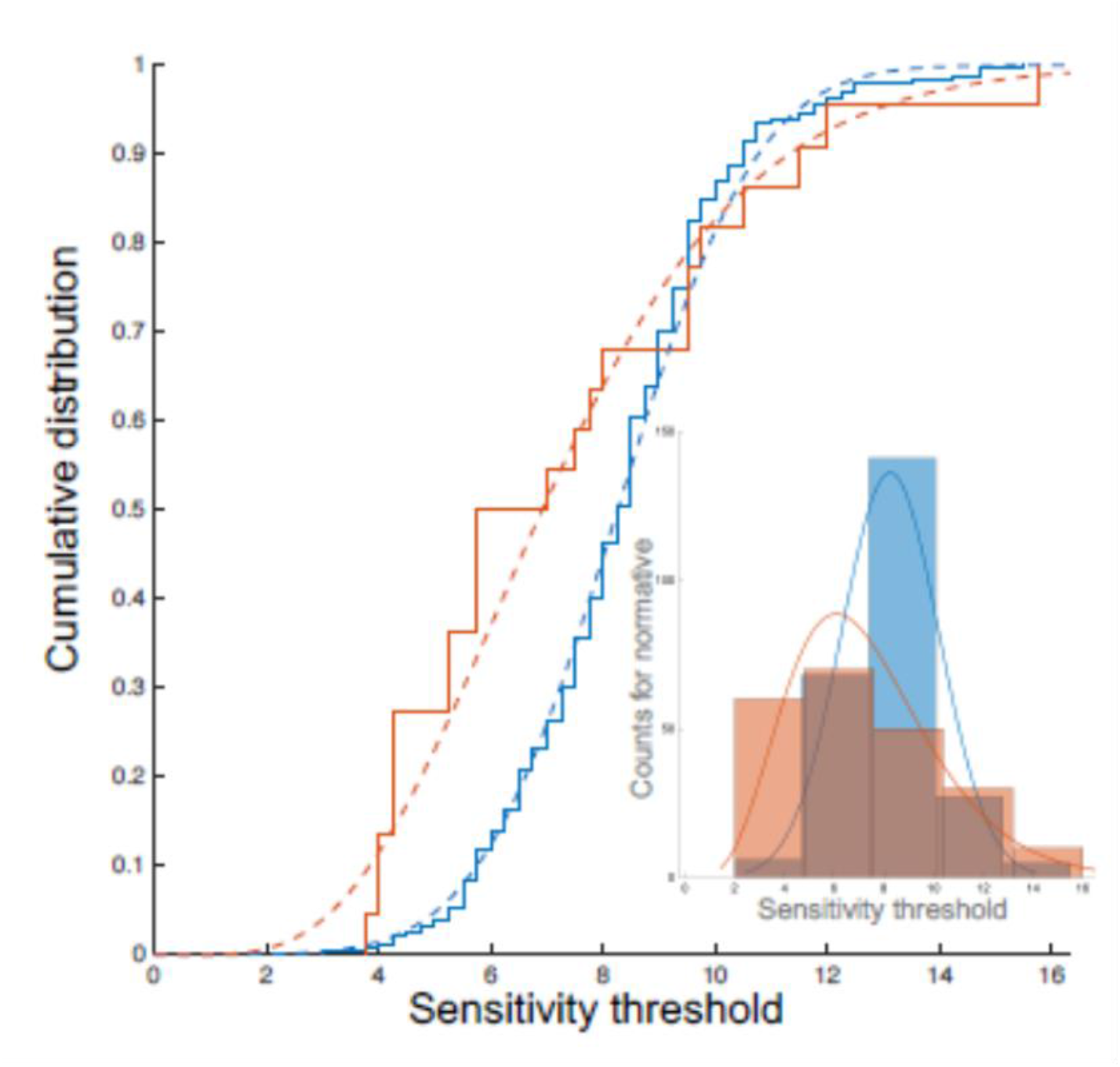
Cumulative distributions of odor detection sensitivity for the normative sample (blue) and Cohort II (orange) which are fit to a gamma distribution (dotted line) and a normal distribution (solid line). Distribution fits were applied as follows: Normative distribution gaussian fit mean=8.26113 [8.01615, 8.50612], sigma=1.95481 [1.79628, 2.14426]; Cohort II Gamma distribution fit mean=4.91205, a=5.99561 [3.37288, 10.6578], b = 1.22059 [0.669847, 2.22415]. Inset: Histograms for odor detection sensitivity for the normative sample (blue; normal fit) and for Cohort II (orange; gamma fit).

### Smell Diagnosis

Categorical diagnoses were examined to define the proportion of students with hyposmia in comparison to our normative sample. From mid-February to March, which included Cohort II and Cohort-IV, the latter assessed between February 14–26, hyposmia was 4-fold more likely than in our normative sample (OR=4.13; CI, 1.8, 9.50). This increased by 7.7-fold for Cohort II (OR=7.74; CI, 3.1, 19.40) and by 13.6-fold for Cohort IIa (OR=13.61; CI, 3.40, 35.66; Fig 3).

**Figure 3.**
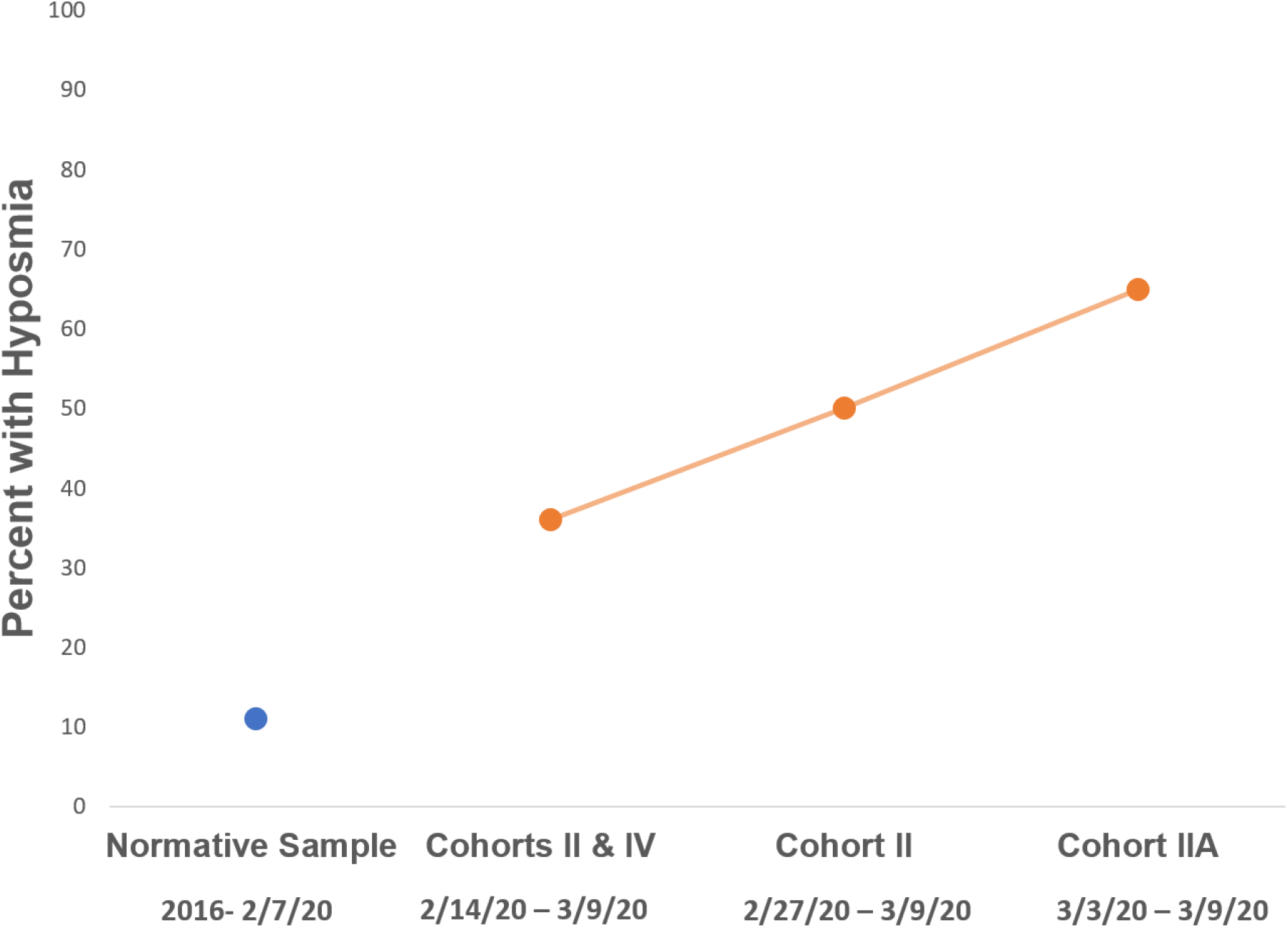
Percentage of students with hyposmia as SARS-CoV-2 was endemic (orange) and the percentage of students with hyposmia in our normative sample. All participants were asymptomatic/pre-symptomatic at the time of assessment.

### Odor Identification and Hedonics

There were no significant differences in odor identification (all P’s >.13) or odor pleasantness ratings (all P’s >.92). Cohort II rated odors as significantly less unpleasant than Cohort III (U=257.0, P<.001, CLES=0.77, CI, 0.51, 1.56). The distribution of unpleasantness ratings for Cohort II was also positively skewed compared to the normative sample (D=0.429, P=.001). Odor detection threshold and unpleasantness ratings were not correlated across the entire sample or within any cohort (all P’s >.16).

### Health Survey

Approximately 15% (N=16) of participants who completed the health survey reported being tested for influenza by clinicians between December 31, 2019 and March 9, 2020. Four had a positive test for influenza (25%), none of whom reported chemosensory changes. Six participants (38%) with negative influenza tests did report chemosensory changes at the time of their flu-like symptoms, consistent with SARS-CoV-2. The last positive influenza test was resulted on February 22, 2020. The only participant formally diagnosed with COVID-19 by a physician reported having anosmia and ageusia two weeks prior other recognized COVID-19 symptoms. Six additional participants suspected they were infected with SARS-CoV-2 but did not obtain testing and four students reported sudden onset anosmia in the absence of other COVID-19 symptoms.

Four Cohort II participants completed the survey, two of whom showed the greatest impairments in odor detection sensitivity (scores of 4 and 4.25). Neither of them reported any subjective change in olfaction, but one experienced nasal congestion and a sore throat a week after olfactory testing, followed by a cough a week later. Another participant (odor detection sensitivity=7) reported the onset of persistent muscle aches almost a month after olfaction testing.

## DISCUSSION

This is the first report of significantly reduced odor detection sensitivity and altered hedonic ratings of odors in asymptomatic students assessed prior to knowledge that the SARS-CoV-2 virus was spreading in the local area. The study employed a comprehensive, psychometrically validated olfaction testing battery administered prior to and during the unique window in which the virus emerged. Our findings suggest that olfactory impairments are the earliest manifestation of SARS-CoV-2 exposure and infection and may therefore be useful for early identification of affected persons to limit contagion.

Notably, these findings were detected in asymptomatic healthy students. The follow-up health questionnaire confirmed that some of the participants who subsequently developed COVID-19 symptoms had the most severe sensitivity decrements yet had no subjective awareness of olfactory impairment. These findings highlight the superiority of psychometric olfaction tests over self-reports for detecting COVID-19, particularly in asymptomatic persons, and strengthen the results of a recent meta-analysis, which found significantly higher rates of COVID-19 related smell loss when objective measures were employed compared to self-report methods [19].

Also noteworthy is that the exposed cohort had intact odor identification, showing spared peripheral olfactory processing despite diminished detection sensitivity. Most studies of COVID-19 patients that have employed objective tests to assess smell identification (but not odor detection) have found significant decrements [20-22]. The only other study to date that has assessed odor detection sensitivity in the context of COVID-19 showed reduced odor detection sensitivity with intact odor discrimination [23], consistent with our findings. Odor identification requires accurate transmission of olfactory stimuli to higher processing regions, indicating intact olfactory receptor neuron function and the fidelity of transmission through the olfactory bulb. Later viral infection and inflammatory pathology may damage the peripheral olfactory pathways, impacting these higher order processes, but our findings suggest that the earliest blunting of smell may have different underpinnings. Further supporting this hypothesis are the results of a recent large-scale multinational study which show two distinct subgroups of COVID-19 patients with olfactory impairment, a group that recovers in under 40 days, and a second group that displays more variable and prolonged recovery [6].

While olfactory receptor neurons do not express the ACE2 receptor and TMPRSS2 that are requisite for viral entry into cells, these are expressed in the epithelial support cells. In particular, ACE2 protein expression is found in the basal layer of the squamous epithelium of the nasal mucosa and nasopharynx [24]. Sensory cells in the nasal mucosa detect viral pathogens and rapidly organize countering immune responses [25] through the activation of nasopharynx-associated lymphoid tissue (NALT). This ancient, innate immune system is the first line of immune defense against airborne pathogens and can activate B and T cell receptor expression.

Humans can also detect sickness in other individuals through an olfactory route, even when they are not ill themselves, activating a specific multisensory threat circuitry that involves the primary and secondary olfactory areas, including entorhinal/piriform cortex, orbitofrontal cortex, inferior frontopolar gyrus, middle frontal gyrus, and thalamus [26]. Activation of this pathway would involve the amygdala, which is consistent with altered perceptions of unpleasantness in the hedonic odor assessments. A resultant reduction of smell and taste sensitivity may be a protective response to pathogen exposure rather than pathology. Along with a shift in the perception of the environment as more unpleasant, behavioral alterations could act to limit social interactions and feeding, preventing exposure to a more pathogenic load of viral particles or transmitting the virus to conspecifics [27]. Social withdrawal is a known component of sickness behavior directed through the central nervous system, but protection against environmental threats through decreasing odor sensitivity is unexplored.

Our findings are consistent with the involvement of some pathways previously associated with immune activation and sickness behavior. It is notable that females and younger persons, who have enhanced chemodetection and immune responses compared to their male and older counterparts, also have better COVID-19 outcomes [28, 29]. Better detection of airborne pathogens may preempt more severe illness though personal immune activation and social withdrawal to prevent viral spread through the social group. Although correlation does not imply causation, a subset of COVID-19 positive individuals with olfactory or gustatory dysfunction appear to have a milder course of illness [7]. Potentially, viral detection in the environment may preclude infection through behavioral and inflammatory mechanisms.

However, without restoration of homeostasis this system could drive inflammation, social dysfunction, and contribute to psychiatric conditions. Our laboratory has shown reduced odor detection sensitivity in association with social withdrawal in males and females with schizophrenia, bipolar disorder, and in a non-clinical sample of persons with olfactory complaints [30-33]. A coordinated immune and behavioral response is not unexpected as infectious diseases account for the greatest amount of human mortality over evolution [34], highlighted again by the COVID-19 pandemic. Viral threats are estimated to underlie a third of mammalian genetic adaptations [35].

The uniqueness of this report rests in the natural experiment wherein SARS-CoV-2 arrived on a campus already having ongoing studies of olfactory function. This research will never be replicated; yet it is not without limitations. The sample sizes of the 2020 cohorts are small, although we still found moderate to large effects and statistical significance. Further, given the few cases of SARS-CoV-2 now known locally at the time Cohort I was tested, it is possible that some Cohort I participants could also have been exposed to the virus prior to testing; we do know that students tested from 2016–2019 were not exposed to the novel virus. Although our subjects were healthy and without nasal symptoms, we did not explicitly assess COVID-19 symptoms at the time of olfaction testing as the studies were not designed to test hypotheses related to infectious diseases. The absence of SARS-CoV-2 positivity confirmation also limits our ability to definitively draw the conclusion about exposure to SARS-CoV-2 at the time Cohort II participated. The retrospective nature of our health survey and lack of local and national access to COVID-19 testing for young adults also limits our health survey results. The State of Ohio, where at almost half of the students reside, only allowed access to COVID-19 testing for non-high-risk persons <60 and non-healthcare workers on June 11, 2020 [36].

### Conclusions

This report adds a scientifically rigorous perspective to the early clinician and patient reports associating anosmia/hyposmia and ageusia/dysgeusia with COVID-19 [37-40], which propelled the American Academy of Otolaryngology to advocate for the CDC and WHO to add these symptoms to COVID-19 screening and urge precautionary isolation for individuals experiencing chemosensory impairment. Crucially, we found that odor detection can be significantly impaired without nasal obstruction or subjective awareness, as participants did not report impaired olfaction at the time of their assessment. While it may not be realistic to administer Sniffin’ Sticks on a population level, daily tracking of odor detection through self-administered protocols could aid in identifying COVID-19 infection in asymptomatic persons and also contribute to our understanding of how olfactory changes may be associated with infection severity.

## Data Availability

We are willing share the data presented in this manuscript.

## ACKNOWLEDGEMENTS

Thank you to the following students for their assistance with data collection, entry, and verification: Brooke Lipnos, Madison Degnan, Rhiannon Gibbs, Wyatt Kaiser, Jenna Kilian, Michael A. Lee, Russell Mach, Maia McLin, Julie Mitchell, Lauren Olson, Piper Sereno, Madeline Scherer, Lisa Stone, Lucia Zook. We also thank Michelle Phares for her assistance in executing online data collection, and Drs. Lee Dixon and Jackson Goodnight for their support in developing the health survey.

